# Genome-wide DNA methylation analysis revealed epigenetic mechanism underlying end stage renal disease

**DOI:** 10.1101/2025.07.20.25331626

**Authors:** Xiaohong Zhou, Dianchun Shi, JinJin Xu, Ling Wang, Resham lal Gurung, Zhiming Ye, M Yiamunaa, Meng Wang, Dongying Fu, Wei Chen, Jinghong Zhao, Hua Gan, Ping Fu, Xiaojun Tan, Jihong Chen, Yaozhong Kong, Haiqiang Zhang, Ming Li, Rajkumar Dorajoo, Xin Jin, Lim Su Chi, Xueqing Yu, Jianjun Liu

## Abstract

End-stage renal disease (ESRD) remains to be a major clinical challenge with persistently high morbidity and mortality, and its molecular mechanisms, particularly those shared among diverse primary kidney diseases during the progression to ESRD, have not been studied. Here we conducted a large-scale two-stage epigenome-wide association study of ESRD in two independent cohorts consisting of 704 controls and 1031 ESRD cases resulting from multiple kidney diseases. We identified 52 ESRD-associated differentially methylated CpG loci (DMLs) that showed consistent association effect between the two cohorts and across diverse kidney diseases. These 52 DMLs implicated 144 candidate genes that showed enrichment in calprotectin complex, RAGE receptor binding and herpes simplex virus 1 infection. Of the 52 DMLs, 5 DMLs were found to be associated with common complications of ESRD, and another 7 DMLs were also found to be associated with renal function decline in early-stage chronic kidney disease, demonstrating their potential as prognostic biomarkers for ESRD risk and related clinical complications. By identifying prognostic biomarkers and revealing the important roles of inflammation and immune dysregulation and renal fibrosis in renal progression to ESRD across diverse primary kidney diseases, our study has contributed greatly to improve clinical management and advance the development of novel therapies for ESRD.

## Introduction

End-stage renal disease (ESRD) poses significant and escalating global health challenges, characterized by substantially high rates of morbidity and mortality^1–3^. Despite its diverse etiologies, ESRD is associated with a high burden of complications, such as anemia, mineral and bone disorders, cardiovascular events^4^, and systemic inflammation, which contribute to its poor prognosis. To reduce the incidence of ESRD, there is a need to understand the molecular mechanisms underlying the renal progression to ESRD and discover biomarkers for early detection of chronic kidney disease (CKD) patients at high risk of progression, thereby enabling timely intervention.

There are multiple factors that can influence the risk of renal progression in kidney diseases, including both environment and genetic factors^5,6^. Common genetic variants linked to CKD progression and renal function decline have been identified through genome-wide association studies (GWAS)^7–9^. In addition, epigenetic modifications, particularly DNA methylation, are increasingly recognized as key contributors to the pathogenesis of kidney diseas^10–13^. Methylated DNA has been regarded as a stable and accessible epigenetic mark for quantitative measurements, making it a suitable source of biomarkers. Recent studies identified DNA methylation changes in blood that were associated with kidney disease and kidney function^13–16^. For example, methylation levels at several CpG sites near *LACTB*, *IRF5*, *ZNF20*, *SLC1A5* and *SLC27A3* genes in blood were significantly associated with estimated glomerular filtration rate (eGFR) and/or risk of CKD^17,18^.

Most epigenome-wide association studies (EWAS) up to date of eGFR and CKD have focused on patients with early-to-mid-stage CKD^16,17,19,20^, limiting insights into the epigenetic mechanisms underlying the severe, intensive and irreversible renal damage in ESRD (Stage 5 CKD). Furthermore, ESRD is a result of renal progression by multiple kidney diseases, such as diabetic kidney disease (DKD), glomerulonephritis, hypertensive kidney disease (HKD) and polycystic kidney disease (PKD), but the previous EWAS of ESRD predominantly focused on diabetic kidney disease^21,22^, which hinders a comprehensive understanding of the common epigenetic mechanisms underlying the late-stage renal progression to ESRD across its diverse etiologies. Notably, ESRD patients experience severe complications, including anemia, mineral disturbances, and accumulation of uremic toxins^23,24^. Mineral metabolism disorders are independently associated with increased cardiovascular mortality and morbidity^25,26^. Therefore, investigating the impact of ESRD-related changes in relation to disease complications may help identify patients at elevated cardiovascular risk.

To overcome the limitations of early studies and advance epigenomic understanding about ESRD, we performed EWAS to examine the variations in DNA methylation levels at 774,631 CpG sites between patients with ESRD (cases) and the ones with prolonged stage 1 CKD for 5 to 10 years (controls) in the Guangdong Provincial People’s Hospital (GDPH) discovery cohort from China. We next performed the validation analysis of the top ESRD DMLs in the independent Khoo Teck Puat Hospital (KTPH) cohort from Singapore. We further explored the effects of the validated DMLs on ESRD-related complications as well as their roles in kidney function decline in early stages of CKD, aiming to identify potential early biomarkers and therapeutic targets for ESRD.

## Results

### Description of Two ESRD cohorts

The current study analyzed two independent ESRD cohorts from China (discovery) and Singapore (validation). The discovery cohort consisted of 196 controls and 802 cases with ESRD from the Guangdong Provincial People’s Hospital (GDPH cohort) in China. The cases were all ESRD patients with eGFR < 15ml/min/1.73m^2^ resulting from five major primary kidney diseases: diabetic kidney disease-ESRD (DKD-ESRD, n = 342), polycystic kidney disease-ESRD (PKD-ESRD, n = 165), IgA nephropathy-ESRD (IgAN-ESRD, n = 144), hypertensive kidney disease-ESRD (HKD-ESRD, n = 81), and lupus nephritis -ESRD (LN-ESRD, n = 70). The controls (n = 196) were patients with CKD stage 1 who preserved renal filtration function of eGFR > 90 ml/min/1.73m² over a 5-10 year period. The validation cohort involved 229 cases with ESRD and 508 controls from the Khoo Teck Puat Hospital (KTPH cohort) in Singapore. The cases were ESRD patients with eGFR < 15ml/min/1.73m^2^ resulting from DKD, and the controls were the patients with a long duration of type 2 diabetes mellitus greater than 10 years with normo-albuminuria and preserved renal filtration function (eGFR > 60 mL/min/1.73 m^2^). Participant baseline characteristics of cases and controls in two cohorts are summarized in Supplementary Table 1.

### Identification of 52 ESRD-associated CpG Loci

The methylation levels at a total of 866,895 CpG sites were profiled in whole blood samples from the GDPH cohort employing the Illumina Infinium MethylationEPIC BeadChip array. The quality control (QC) analysis led to the exclusion of 2 samples due to sex swaps, as well as 92,264 low-quality CpG probes (e.g. non-specific, cross-hybridizing, or high-impurity probes identified via the greedycut algorithm). This resulted in a dataset of 774,631 CpG probes across 998 individuals (802 cases and 196 controls) for downstream analysis in the GDPH cohort. In the KTPH cohort, there were 866,895 CpG probes successfully assayed, and 888,78 CpG probes were excluded following the same probe-level QC procedures as in the GDPH cohort, and no samples failed quality control, leaving 737 samples (229 cases and 508 controls) and 778,017 CpGs for analysis.

As the discovery analysis, we first conducted EWAS using 196 controls and each of the five ESRD case groups with different primary kidney diseases (DKD, PKD, IgAN, HKD and LN). The CPACOR pipeline was employed to adjust for the effects of technical and biological confounders on DNA methylation level measurements. Principal component analysis (PCA) was performed using the confounder-adjusted DNA methylation level measurements, which demonstrated a good match between all ESRD cases and controls (Supplementary Figure 1). The confounder-adjusted DNA methylation level measurements were then used for association analysis. We minimized the inflation effect of association results using the Bacon method^27^, which is a Bayesian method for estimating the empirical null distribution to control for the bias and inflation of the statistical results from EWAS. We observed no or negligible inflation of the summary statistics in all five EWAS (DKD-ESRD inflation= 0.94, PKD-ESRD inflation= 1.1, IgAN-ESRD inflation=1.06, HKD-ESRD inflation= 1.03, LN-ESRD inflation= 1.1, Supplementary Figure 2). The EWAS results for each ESRD group are summarized in Supplementary Table 2 and Figure 1b-f.

**Figure 1.**
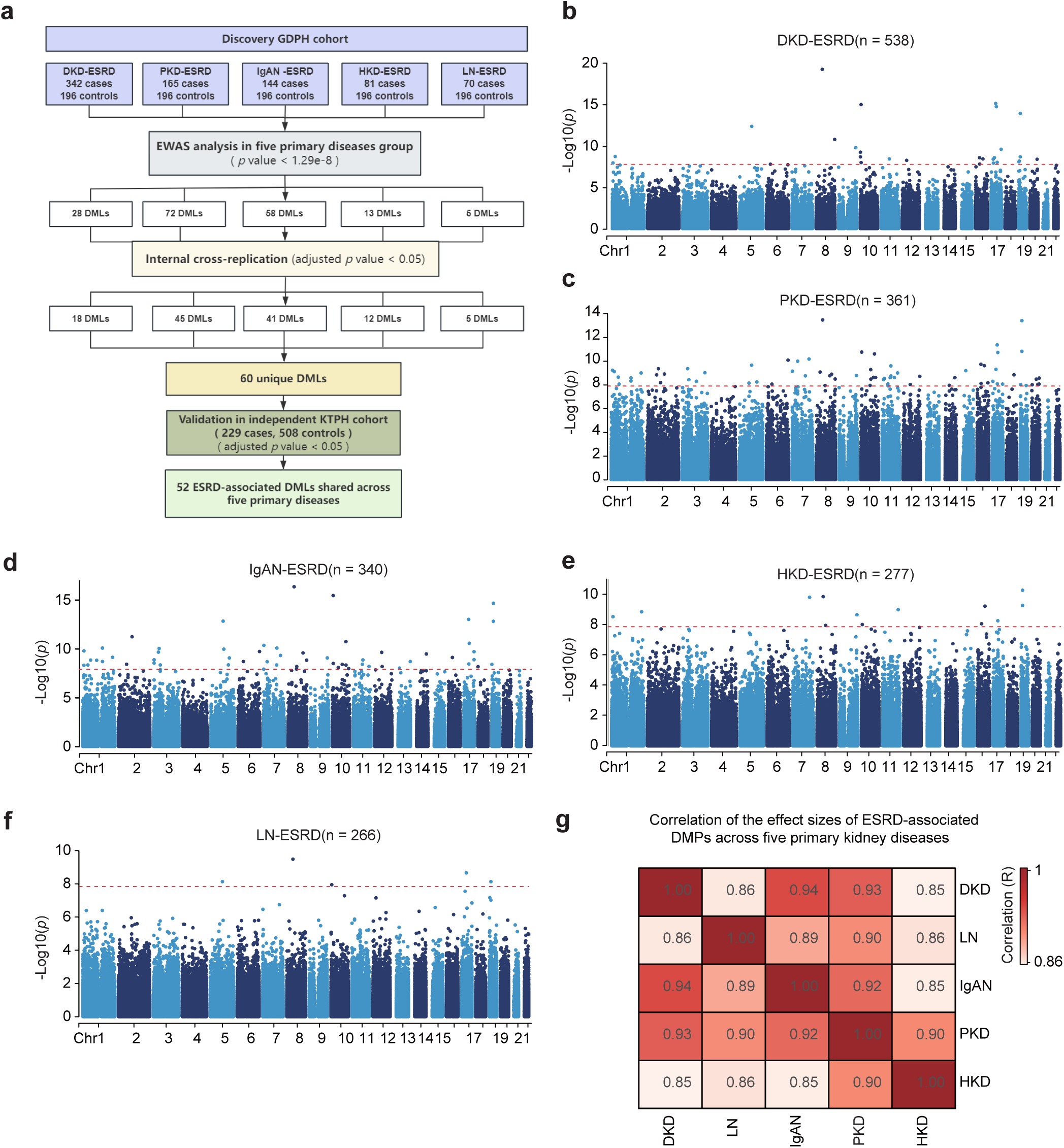
Identification of common ESRD-associated differentially methylated loci (DMLs) across five primary kidney diseases. **(a)** The study design for identifying ESRD DMLs across five primary kidney diseases. **(b-f)** Manhattan plots for the EWAS results for ESRDs caused by DKD, PKD, IgAN, HKD and LN respectively. The red dash line shows the genome-wide significance threshold (1.3e-8). Chromosome and genomic coordinates are shown on the x-axis, with the two-sided statistical significance (–log₁₀ *P*) values on the y-axis. **(g)** Heatmap of pairwise Spearman correlation coefficients among ESRD primary disease-specific effect size estimates of EWAS across 60 common ESRD DMLs.

Aiming to discover DNA methylation changes that are shared among the ESRD patients with five primary kidney diseases, we compared the results from the five EWAS analyses and identified 60 DMLs whose methylation levels showed genome-wide significance (*P* < 1.3 × 10⁻⁸, see the Methods) in at least one ESRD group and also Bonferroni-corrected *P* < 0.05 in the other groups, (Figure 1a). We further performed a validation analysis of these 60 shared DMLs in an independent cohort from Singapore (KTPH cohort, n=737) and confirmed that 52 of the 60 DMLs showed consistent associations (Bonferroni-corrected *P* < 0.05, Supplementary Table 3), with effect sizes highly correlated between discovery and validation cohort (Spearman r = 0.78–0.90; Supplementary Table 3, Supplementary Figure 3).

Across five ESRD groups, the effect sizes of these 52 DMLs showed high consistency, with r² values exceeding 0.7 in all pairwise comparisons, indicating they reflect common ESRD pathogenic mechanisms regardless of the primary disease (Figure 1g). In particular, nine of these DMLs achieved genome-wide significance across at least three ESRD groups with different primary diseases (Supplementary Figure4). Notably, the cg17944885, located in the intergenic region between genes *ZNF788P* and *ZNF625-ZNF20*, showed the genome-wide significant association in all the five ESRD groups and another 2 DMLs, cg13241457 located in the promoter of *C16orf54* and cg25544931 located between *ZNF788* and *ZNF20*, achieved genome-wide significance across four ESRD groups.

We further evaluated whether the 52 ESRD-associated DMLs identified in our study were enriched among the CpG sites previously reported to be linked with other diseases and related traits, by querying two publicly available EWAS repositories (EWAS atlas and the MRC-IEU EWAS Catalog). As expected, the ESRD DMLs showed the strongest enrichment in the EWAS results for eGFR and CKD, being 121- and 54-fold more likely than expected by chance to overlap with the CpG sites associated with eGFR and CKD, respectively (Bonferroni-corrected *P* < 0.05; Supplementary Figure 5). These findings confirmed that the 52 ESRD-associated DMLs and related CpG sites reflect shared epigenetic regulatory mechanisms involved in impaired renal function.

To assess whether the significant associations at the 52 DMLs were influenced by the clinical factors that are known to be associated with kidney disease progression, a sensitivity analysis was conducted in the GDPH cohort. The association analysis was performed in the regression model that was adjusted for each of the baseline clinical factors that are significantly different between ESRD cases and controls in the GDPH cohort, including total cholesterol (TC), low-density lipoprotein cholesterol (LDLC), triglycerides (TG), high-density lipoprotein cholesterol (HDLC) and uric acid (UA). Remarkably, the sensitivity analysis indicated that the association results for the 52 DMLs were robust and not influenced by these clinical factors (Supplementary Figure 6), as evidenced by the high correlation between the effect sizes from the original (without adjustment for various factors) and sensitivity model (with adjustment for various factors).

### Impact of ESRD DMLs On Clinically Common Complications of ESRD

We also investigated the impact of the 52 DMLs on the clinical complications of ESRD in the GDPH cohort, including uncontrolled anemia (< 115 g/L)^28^ and chronic kidney disease-mineral and bone disorder (CKD-MBD). CKD-MBD manifests as markedly high parathyroid hormone (PTH) levels, hypocalcemia and hyperphosphatemia in the late stages of CKD (KDOQI Clinical Practice Guidelines, 2017), all of which were known to significantly increase both cardiovascular (CV) and all-cause mortality risks in ESRD patients^29–31^.

Through an ESRD case-only analysis, we estimated the association of these 52 DMLs with these complications. Of the 52 DMLs, five were associated with ESRD-related complications (FDR < 0.05): cg07054804 (*MRVI1*) and cg16980393 with elevated PTH levels; cg26171235 with hyperphosphatemia; cg13854688 with hypocalcemia; and cg19526450 (*ARHGAP26*) with uncontrolled anemia (Supplementary Table 4). Noticeably, the methylation level at cg07054804, located in *MRVI1*, was correlated with the gene expressions of nine genes in blood, including *NDRG2, DLEU7* and *CD93*^32^. Among these genes associated with cg07054804, the genetic polymorphisms or altered expressions of *DLEU7* and *CD93* were previously linked to decreased eGFR^11,33^, CVD risk^34^, and reduced bone mineral density^35,36^, suggesting that these ESRD-related DMLs may play significant roles in the development of ESRD-associated complications. In addition, we compared the effects of these 52 DMLs on the complications with the ones on ESRD itself and found that the effects of the 52 DMLs on CKD-MBD markers were significantly and positively correlated with their effects on ESRD (Figure 2).

**Figure 2.**
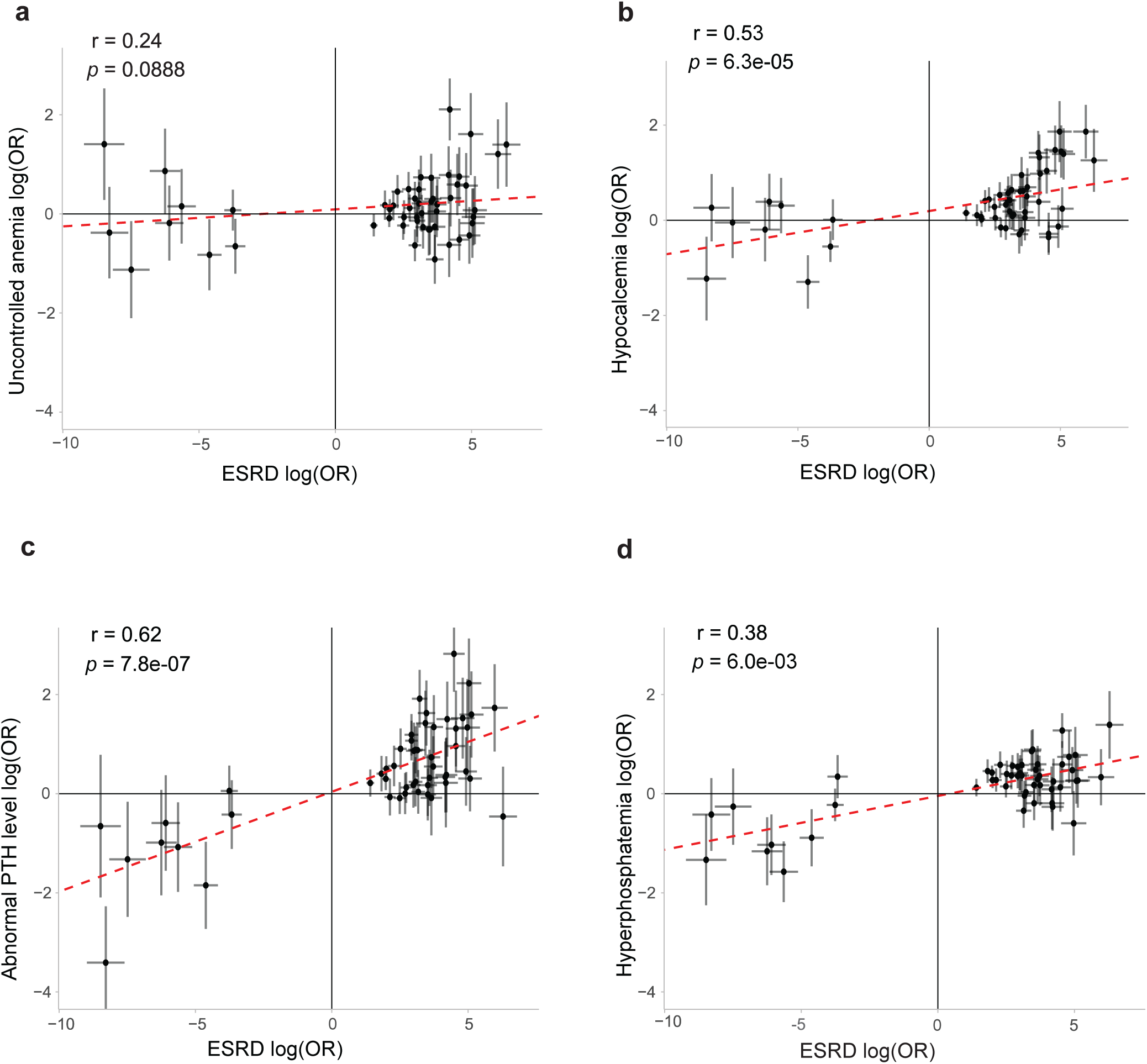
Correlation analysis of the estimated effect sizes of EWAS between ESRD and related-complications for 52 ESRD DMLs. Comparison of effect size estimates between ESRD and ESRD-related complications **(a)** uncontrolled anemia, **(b)** hypocalcemia, **(c)** hyperparathyroidism, and **(d)** hyperphosphatemia. The y-axis displays log odds ratios (logORs) from logistic regression analyses evaluating associations between 52 DMLs and the complications among ESRD patients. The x-axis presents corresponding logORs derived from the association analysis of ESRD groups, each defined by a distinct primary kidney disease. Spearman’s rank correlation (two-sided) was applied to assess the concordance between the two sets of logORs. The red dashed line illustrates the fitted linear trend. Error bars denote 95% confidence intervals.

### Impact of ESRD DMLs on Renal Function Decline in Early CKD

Aiming to identify epigenetic biomarkers for predicting CKD progression to ESRD, we investigated the roles of the 52 ESRD DMLs in kidney renal function decline in early-stage CKD. In particular, we examined their impact on key renal function markers, including eGFR based on serum creatinine (eGFRcrea), eGFR based on combined serum cystatin C and creatinine (eGFRcys-crea), and blood urea nitrogen (BUN) in early-stage CKD by evaluating: (1) their associations with eGFRcrea using published EWAS summary data, and (2) their overlapping with the GWAS loci for eGFRcys-crea and BUN using colocalization and Mendelian randomization (MR) analyses that have been commonly utilized by previous studies to investigate the role of DMLs in complex traits^18,37–40^ to link DNA methylation levels with complex traits.

We performed meQTL analysis of the 52 ESRD-associated DMLs in the GDPH cohort where both genome-wide SNP genotyping and DNA methylation data were available, to identify genetic variants that influence methylation levels (meQTL SNPs) at DMLs. A total of 26 ESRD DMLs were significantly associated with at least one meQTL SNP (P < 5 × 10⁻⁸; Supplementary Tables 5–7). The top three cis-meQTL loci for cg00037671, cg09299075, and cg00829404 (Figure 3a,b and d) and the top two trans-meQTL loci for cg24799186 and cg17124293 (Figure 3c and e) were all supported by clusters of SNPs that are in moderate to high linkage disequilibrium at each locus (indicated by the purple diamond in Figure 3).

**Figure 3.**
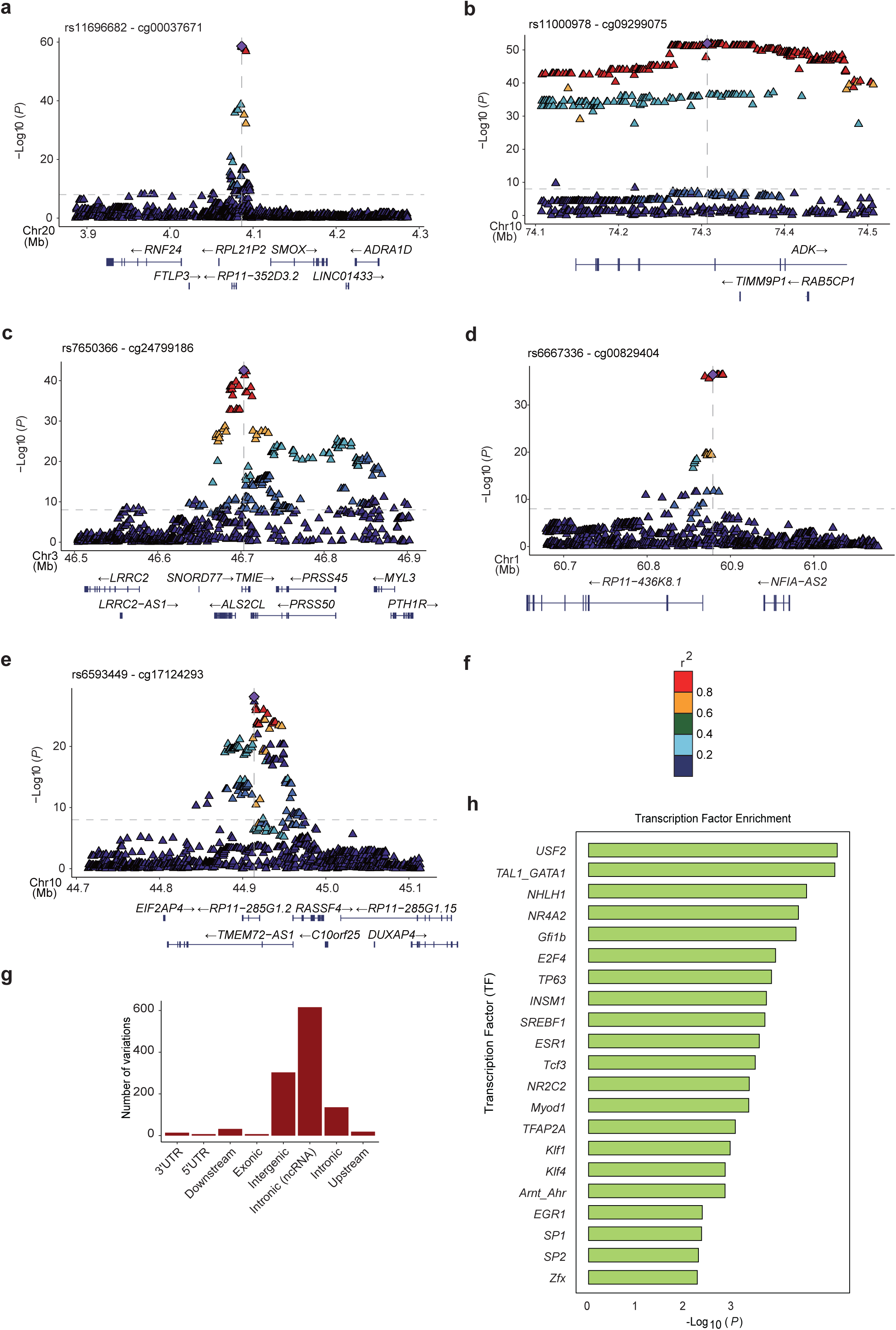
meQTL analysis of ESRD DMLs. For each of 52 ESRD DMLs, meQTL analysis was conducted using DNA methylation data and available genotypes from GDPH participants (n = 924). **(a–e)** Regional association plots (–log₁₀*P*) for top five significant meQTL loci. Each diamond represents a SNP, with the most significantly associated SNP shown in purple and labeled by rsID. **(f)** SNP colors indicate linkage disequilibrium (LD), computed using genotype data from 3,884 Chinese individuals, between the top SNP and surrounding variants. **(g)** Genomic distribution of 3,320 cis-meQTL SNPs. **(h)** Enrichment of cis-meQTL SNPs in transcription factor motifs, which was assessed by binomial testing.

The cis-meQTL SNPs for the ESRD DMLs are predominantly located in intergenic and intronic regions, particularly the ones of ncRNAs (Figure 3f). We observed that 21 transcription factor binding sites (TFBS) are enriched around the cis-meQTL SNPs (Figure 3g, Supplementary Table 8), some of which are known to influence DNA methylation by recruiting DNA methyltransferases (DNMTs) or ten-eleven translocation enzymes (TETs) near their binding sites^41–45^, or are linked to kidney disease processes like inflammation and fibrosis^46–50^. These findings provide strong support for the role of meQTL SNPs in regulating DNA methylation at the ESRD-associated DMLs.

We next evaluated the relationship between DMLs and three renal function markers. For eGFRcrea, we identified and evaluated 23 DMLs with available summary statistics in existing EWAS data for eGFRcrea^22,51^. Among 23DMLs, 9 DMLs (39.13%) were found to show significant association with inversed effect directions compared to their associations with ESRD (Supplementary Table 9). For eGFRcys-crea and BUN, we performed the colocalization analyses and Mendelian randomization using the meQTL SNPs of the ESRD DMLs. Among 26 ESRD DMLs associated with at least one meQTL (association *P* < 5 × 10^-8^), 9 DMLs (34.62%) showed genetic colocalization with eGFRcys-crea and/or BUN (Coloc Posterior Probability [Coloc PP.H3 or Coloc PP.H4] ≥ 0.6) GWAS loci (Supplementary Table 10). Among 41 ESRD DMLs with at least one cis-genetic instrument at a suggestive significance (*P* < 5 × 10⁻⁴; r² of LD < 0.2; Supplementary Table 11), we found evidence supporting significant causal effects of DNA methylation at 6 ESRD DMLs (14.63%) on eGFRcys-crea and/or BUN levels (Supplementary Table 12).

By applying a set of complementary methods, 17 of the 52 DMLs (32.69%) showed associations with at least one kidney function marker, and 7 DMLs were associated with at least two renal function markers (Supplementary Table 13), including two that are associated with all three kidney function traits (eGFRcrea, eGFRcys-crea, and BUN). For example, cg04816311, located within *C7orf50*, exhibited a significant association (*P* = 2.88e-8) with eGFRcrea, and the cg04816311-related meQTL was colocalized with the loci associated with both eGFRcys-crea and BUN (Coloc PP.H3 > 0.99). Cg09299075 near *ADK*, was associated with eGFRcrea in external EWAS (*P*=7.8e-6), and shared same genetic variants with both BUN and eGFRcys-crea (Coloc.PP.H3 or Coloc.PP.H4 > 0.7). Notably, genes linked to these methylation sites (Supplementary Table 13), such as *PDIA3* (*ERP57)*^52^, *AFF3*^19,53^, *S100A8*^54^, *S100A9*^54^, *C7orf50* ^55–57^ and *RPTOR*^58^, have been reported to be linked with kidney injury, kidney fibrosis and the accelerated progression of kidney disease. Together, these seven CpG sites may serve as early epigenetic markers to identify individuals with early-stage CKD who are at higher risk of progressing to ESRD.

### Gene-annotation and Enrichment Analyses

The majority of the ESRD DMLs are located in the gene body (51.9%) (Figure 4a), where DNA methylation is increasingly recognized as a key regulator of gene expression, with significant implications for various biological processes^59,60^. To gain biological insight into ESRD DML-related molecular mechanisms, we identified ESRD DML-related candidate genes using four criteria (Supplementary Table 14): (1) genes harboring DMLs within coding regions, (2) genes regulated by DML-associated active enhancers in blood, (3) genes whose expression correlates with ESRD DML methylation levels in blood or kidney, and (4) genes with cis expression quantitative trait loci (cis-eQTLs) co-localized with cis-meQTLs (see Methods). In total we have identified 144 candidate genes for 49 ESRD DMLs (49/52, 94%) (Supplementary Table 16). Figure 4b and Supplementary Figure 6 illustrated the number of candidate genes annotated by each approach. Among the 144 annotated genes, 25 were influenced by active enhancers containing DMLs, highlighting the role of enhancer-associated DNA methylation in ESRD-related gene expression regulation. Furthermore, eQTM database analysis revealed that the expression levels of 15 genes in the kidney and 32 genes in blood are highly associated with DNA methylation at ESRD-associated DMLs. And 57 unique genes were found to potentially colocalize with a nearby ESRD DML in blood, indicating the presence of shared causal variants affecting both gene expression and DNA methylation patterns (Supplementary Tables 15). Notably, the expression levels of *DLEU7* and *SP140* are uniquely regulated by eight and six DMLs, respectively, underscoring the complex, multi-DML regulatory mechanisms involved in ESRD pathogenesis.

**Figure 4.**
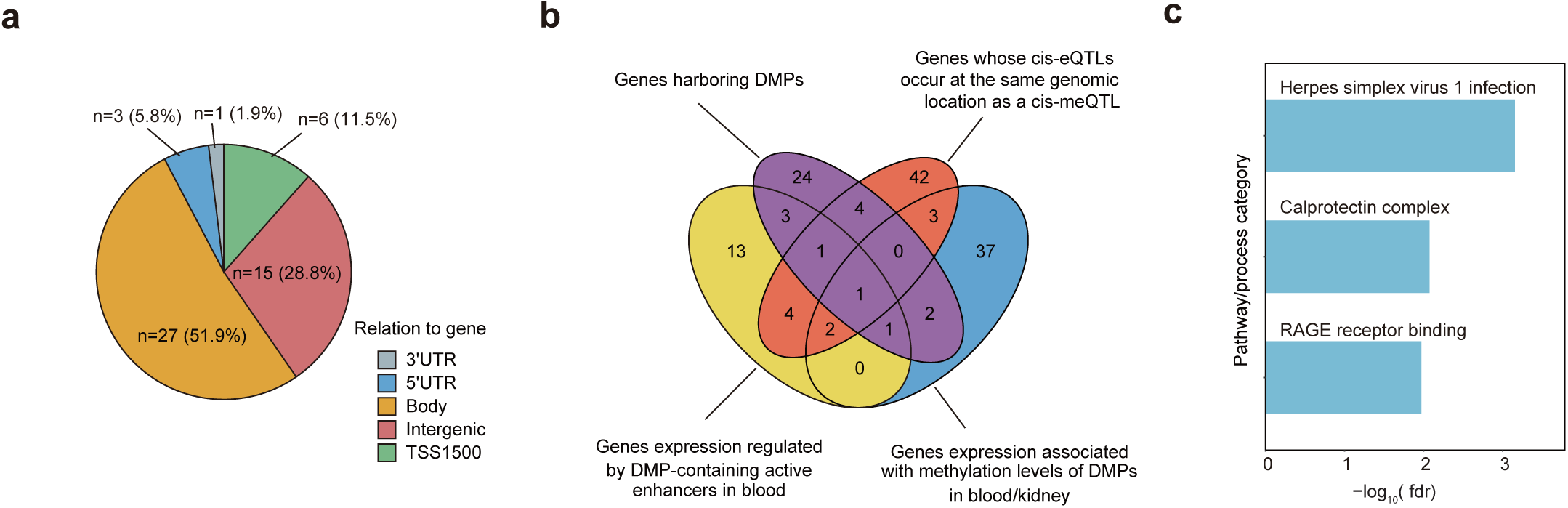
Genes annotated to ESRD-associated DMLs and gene set enrichment analysis. **(a)** Genomic distribution of 52 ESRD-associated DMLs. **(b)** Venn diagram showing genes annotated to methylated sites, categorized into three groups: genes harboring DMLs, genes regulated by DML-containing active enhancers in blood, genes whose expression is associated with DML methylation levels in blood/kidney, and genes whose cis-eQTLs occur at the same genomic location as a cis-meQTL. Overlapping regions indicate shared genes. **(c)** Enriched biological processes/pathways associated with ESRD-related DMLs, identified using a FDR threshold of 0.05.

We then conducted gene set enrichment analysis (GSEA) using the 144 candidate genes revealed by gene annotation analysis. The GSEA identified three enriched processes and pathways (Figure 2 d) involved in inflammatory responses and kidney fibrosis, including herpes simplex virus 1 infection (FDR = 6.68e-4, KEGG: 05168), calprotectin complex (FDR = 0.008, GO:1990660), and receptor for advanced glycation end-products (RAGE) receptor binding (FDR = 0.01, GO:0050786). Notably, both RAGE receptor binding or Calprotectin complex pathways include *S100A8* and *S100A9*, suggesting their pivotal regulatory roles in inflammation, immune dysregulation associated with ESRD (Supplementary Table 17).

## Discussion

In the current study, we performed a two-stage EWAS in two independent ESRD cohorts and discovered 52 CpG loci (DMLs) whose differential methylation levels are strongly associated with ESRD, showing consistent effects among five major kidney diseases, including DKD, PKD, IgA N, HKD and LN. By integrating both genomic and epigenomic data, we were able to link these 52 DMLs to 144 candidate genes and found the enrichment of these candidate genes in three pathways pivotal to end-stage renal pathology in kidney diseases: (i) herpes-simplex-virus-1 infection, driving inflammation and immune dysregulation; (ii) RAGE receptor binding; and (iii) the calprotectin complex, both of which promote renal fibrosis and chronic kidney inflammation. Crucially, five DMLs showed significant associations with key ESRD complications, including anemia and CKD-MBD, suggesting their potential utility in the identification of ESRD patients at high risk for complications. In addition, seven DMLs were also found to be associated with decreased kidney functions in early-stage CKD, highlighting their potential as prognostic biomarkers for progression to ESRD in CKD patients.

Earlier EWAS of kidney function traits mainly focus on early- or/and middle-stage CKD^17,19,20,22^. There were a few EWAS on ESRD, but these efforts were only focused on ESRD of specific kidney disease, such as T1D where the biological insights were likely to be T1D specific^21,61^. These early studies, therefore, provided limited insights for the late-stage progression to ESRD, the severe and irreversible endpoint of renal function decline for all the major kidney diseases. By analyzing the ESRD cohorts resulting from five major metabolic and inflammatory kidney diseases, our study has overcome the limitations of previous studies and provided novel insight into epigenomic mechanisms underlying the late-stage renal progression to ESRD that are likely shared among different kidney diseases. For instance, three ESRD-related DMLs—cg17944885 (located near *ZNF788*), cg25544931 (between *ZNF763* and *ZNF433-AS1*), and cg13241457 (in the promoter region of *C16orf54*), showed genome-wide significant association in at least four patient groups with distinct primary kidney diseases. Supporting our results, abnormal methylation levels at cg17944885 have been significantly associated with multiple kidney traits, including lower eGFR levels, DKD, and renal fibrosis in micro-dissected human tubulointerstitial compartments^17,22,51^. Notably, prioritized genes corresponding to these three CpGs, including *ZNF20, ZNF700, ZNF763, ZNF136, ZNF439* and *ZNF44,* are enriched in the herpes simplex virus 1 (HSV-1) infection pathway. Although direct evidence linking HSV-1 infection to CKD progression is limited, this observation aligns with prior studies showing that infection with a related Herpesviridae member, herpes zoster (HZ) infection, is associated with the progression of CKD to ESRD^62^, and the increased mortality risk in ESRD^63^. Patients with CKD and ESRD exhibit immune dysfunction, rendering them more susceptible to infections caused by both HSV-1 and HZ^64,65^. We hypothesized that these infections may contribute to kidney damage via potential mechanisms involving proinflammatory cytokine production, chronic systemic inflammation, and immune dysregulation.

In addition to HSV-1 infection, our analysis also revealed two addition pathways linked to most severe renal damage, including RAGE receptor binding ^66^ (*S100A12, S100A8,* and *S100A9*) and calprotectin complex ^67,68^ (*S100A8,* and *S100A9*). Calprotectin, a protein complex assembled from S100A8 and S100A9, both EF-hand calcium-binding proteins, serves as a damage-associated molecular pattern that activates the receptor for RAGE and toll-like receptor 4. Through this activation, calprotectin contributes to the progression of renal damage, fibrosis, and cardiovascular diseases.^54,69–71^. S100A8/A9 knockout (−/−) mice were protected from renal interstitial fibrosis in diabetic nephropathy and obstructive nephropathy^54,71,72^. Higher circulating levels of S100A8/A9 have been identified as robust and independent predictors of vascular calcification, adverse cardiovascular events, and mortality among patients with chronic kidney disease ^67^. In addition to S100A8/A9, S100A2 knockout also attenuated interstitial fibrosis in kidneys via suppression of FoxO1-mediated epithelial-mesenchymal transition^73^. Clinically elevated S100A12 levels predict rapid CKD progression (≥50% eGFR decline/ESRD over 10 years) and correlate with systemic inflammation, complications, and increased mortality in ESRD patients^74–76^. Moreover, plasma S100A12 concentrations have also been found to be significantly associated with CVD prevalence in patients undergoing hemodialysis, and the S100A12/RAGE axis might represent a key mediator connecting inflammatory processes with atherosclerosis and vascular calcification^76,77^. Our study has suggested a potential role for S100A12, S100A8, and S100A9 in the development of ESRD and its associated cardiovascular and metabolic disorders.

Our study suggests that ESRD-associated DNA methylation loci may serve as potential biomarkers for predicting the risk of ESRD-related clinical complications. Specifically, our case-only analysis has identified five ESRD-associated DMLs that also showed strong association with CKD-MBD in ESRD patients, such as cg07054804, located near *MRVI1*, which was linked to elevated PTH levels. As a central feature of CKD-MBD, elevated PTH promotes vascular calcification and cardiovascular morbidity in advanced CKD/ESRD, mechanistically connecting mineral-bone dysregulation to elevated cardiovascular risk. This association is mechanistically reinforced by the biological context of cg07054804 (*MRVI1*). The *MRVI1* gene has been shown to be significantly associated with coronary artery disease^78^, unstable angina^79^ and lacunar stroke^80^, of which encoded protein plays an important role in intracellular calcium release, vascular inflammation and platelet adhesion^80,81^. Interestingly, the methylation level of cg07054804 was associated with differential expression of *NDRG2*, *DLEU7* and *CD93* in blood^32^. *NDRG2* mitigates renal fibrosis by suppressing TGF-β1/Smad3 signaling in tubular epithelial cells^82^, regulates bone remodeling through osteoblast modulation^83^, and protects against cardiovascular damage by reducing cardiomyocyte apoptosis and improving myocardial remodeling in heart failure^84^. Moreover, although *CD93* and *DLEU7* have limited associations with CKD–MBD, polymorphisms in both genes are genetically linked to coronary artery disease^34,85^. The methylation at cg07054804 may influence calcium PTH levels by regulating the expression or function of *MRVI1*, *NDRG2*, *DLEU7* and *CD93*, thereby potentially contributing to the increased risk of cardiovascular disease in ESRD patients. Our results could provide evidence to clarify the potential epigenetic mechanism of ESRD-associated complication, warranting further functional and clinical investigation.

By integrating EWAS summary statistics with GWAS colocalization and Mendelian randomization analyses, we identified seven ESRD-associated DMLs linked to decreased kidney function in cohorts comprising early CKD patients and healthy individuals. Their presence in early-stage disease establishes them as potential biomarkers for predicting ESRD risk in early CKD. The *AFF3* locus (cg22386583) demonstrates strong evidence for its role in CKD progression: Its methylation associated with eGFR decline in early stage ^22^, and a nearby variant (rs7583877) is linked to ESRD in type 1 diabetes ^53^. *AFF3* promotes renal fibrosis via TGF-β-driven fibroblast activation and ECM deposition, suggesting its potential as a predictor of fibrosis-driven CKD progression ^53^. Another significant marker, *ERP57* (cg06158227), is involved in renal fibrosis, which functions as an early indicator of kidney injury and plays a continuous role in CKD progression^52^. Animal studies show that elevated transglutaminase 2 (TG2) activity is essential for nephropathy and renal scarring. In this context, increased ERP57 export in renal fibrosis may compensate by attenuating aberrantly activated TG2 in the extracellular matrix^86^. Nevertheless, validation through prospective cohort studies and predictive modeling remains essential to confirm their clinical applicability across the full spectrum of CKD.

In summary, our study has expanded the biological understanding about the molecular mechanisms underlying ESRD, particularly the epigenetic mechanisms involved in the late-stage renal progression to ESRD that are shared among different primary kidney diseases. Particularly, our study highlighted the critical role of dysregulated immune and inflammatory responses and renal fibrosis in ESRD, involving genes such as *S100A8, S100A9* and *S100A12,* and other genes which are associated with the pathways of HSV-1 infection, RAGE receptor binding, and the calprotectin complex. Further functional and clinical studies of these pathways may provide novel targets for therapeutic development. Our study has also identified several epigenetic biomarkers with good potential for identifying early-stage CKD patients with high risk for ESRD and thus preventing renal failure through early intervention, as well as ESRD patients with high risk for complications for informing personalized, complication-focused treatment strategies.

## Methods

### Discover cohort: GDPH study

A study in China consisted of 802 ESRD patients in five primary kidney diseases and 196 disease controls. Primary causes of ESRD included DKD (n = 342), PKD (n = 165), IgAN (n = 144), HKD (n = 81) and LN (n = 70). ESRD was identified using diagnostic codes from either ICD-9-CM (585) or ICD-10-CM (N18.5/N18.6), and included individuals with a history of kidney transplantation. This condition reflects a permanent and severe loss of kidney function, defined by a eGFR persistently below 15 ml/min/1.73 m², clinical evidence of uremia, and the requirement for long-term renal replacement therapy. Disease controls were CKD patients with a disease duration of 5–10 years who consistently maintained an eGFR > 90 ml/min/1.73 m² and remained in CKD stage 1 throughout the entire period. CKD was characterized by ≥ 3-month persistent nephropathy involving structural/functional derangements consequential to health .^87^

During baseline assessment, standardized interviews and clinical examinations were administered to all enrolled participants, with collection of blood samples. Detailed interviews were conducted to collect information on age, sex, medication history, and levels of hemoglobin (Hb), albumin, calcium, phosphorus, parathyroid hormone (PTH), and blood lipids.

In compliance with the Declaration of Helsinki, the Institutional Review Board of Guangdong Provincial People’s Hospital authorized this investigation, for which all subjects provided documented informed consent. EWAS was performed using genomic DNA extracted from peripheral blood.

### Validation cohort: KTPH study

The KTPH study is a case-control study involving 737 individuals, including 229 ESRD cases attributed to DKD and 508 controls. Controls were selected from individuals with type 2 diabetes for more than 10 years who did not develop DKD. Baseline demographic of ESRD patients were collected at enrollment. All participants provided written informed consent compliant with institutional ethics protocols. Multidimensional demographic characteristics and clinical phenotypes were systematically compiled within the electronic health repository. Genomic DNA from peripheral blood mononuclear cells were used for EWAS

### Blood DNA Methylation

For GDPH cohort, We utilized the Illumina Infinium MethylationEPIC array to profile DNA methylation across the genome. QC and preprocessing steps for DNA methylation measurements were executed with the R package RnBeads (version 2.12.2)^88^. In the quality control step, 17,371 sites overlapping with SNPs were removed due to their potential to map to multiple genomic locations. Additionally, 43,463 non-specific probes or probes with a high likelihood of cross-hybridization were removed. 12,812 probes with the highest impurity were removed based on the Greedycut algorithm (detection p < 0.01). In each iteration, the algorithm generated a matrix of retained and removed measurements, calculated the false positive rate (α) and sensitivity (s), and selected the values that maximized the expression “s + 1 – α”, thereby giving equal weight to both sensitivity and specificity. Probe-type bias correction was implemented through beta-mixture quantile (BMIQ) normalization, a gold-standard method for epigenetic array data processing as established in prior methodology studies^89^. We further removed 1,293 probes with specified contexts (e.g., non-CpG), 17,119 probes located on sex chromosomes, and 206 probes with a missing rate greater than 10%. No participants were excluded due to a probe missing rate exceeding 5% (call rate ≥ 95%). Imputation of the beta methylation matrix was performed using the “mean” method in the RnBeads package^88,90^.

The quality control (QC) analysis led to the exclusion of 2 samples due to sex mismatches. All GDPH cohort samples met the 95% CpG call rate quality threshold. The final methylation working dataset comprised 774,631 CpG methylated probes for 998 samples. M-values were computed from the normalized beta values using the following formula: M = log₂(β / (1 − β)), where β denotes the methylation beta value.

In KTPH cohort, DNA methylation at 866,895 CpGs were also profiled using the EPIC array. Quality control and data pre-processing steps were performed using the same procedures as in the GDPH cohort. 43,463 cross-hybridizing probes and 17,371 SNP-proximal sites (±3 bp) were filtered out during quality control. A total of 9,360 unreliable probes were removed using the Greedycut algorithm (p < 0.01), while no samples were excluded. An additional 17,198 probes located on sex chromosomes, 1,313 probes with specified contexts, and 173 probes with a call rate below 95% were further removed. All samples passed the QC based on the Greedycut algorithm and had a call rate > 95%. Finally, 737 samples with 778,071 CpG sites passed all strict QC procedures.

### Technical factors

The Illumina EPIC array includes a set of internal control probes designed to monitor various sample preparation steps (bisulfite conversion, hybridization, etc.) To capture and adjust for technical variation and batch effects, we performed principal component analysis (PCA) on the non-negative intensity values of these internal control probes. DNA methylation analyses were subsequently adjusted for the top 30 principal components derived from this control probe PCA.

### Blood cell-type subpopulations

In both GDPH and KTPH cohort, we used EpiDISH cell subtype deconvolution approach^91^ (version 2.10.0) to estimate fractional abundances of six peripheral immune subsets: neutrophils, monocytes, B-cells, CD4⁺ T-cells, CD8⁺ T-cells, and NK cells. Reference panels were sourced from “centDHSbloodDMC.m”.

### Smoking score

DNA methylation status is markedly affected by smoking. Smoking status were predicted for all individuals using the “Epismoker” R packages (version 0.1.0).^92^

### Association analysis

In both the GDPH and KTPH cohorts, a three-step procedure incorporating linear and logistic regression models was implemented to eliminate confounding factors and elucidate the correlation between ESRD and DNA methylation at each CpG site. Notably, M-values (defined as 𝑙2(/1−)) instead of β-values was used on EWAS

***1)*** For each CpG site in the cohort (n=998), we performed covariate-adjusted linear regression with: biological variables (sex, age, smoking), estimated immune cell fractions, and control probe-based PCs 1-30

***DNAme ∼ Age + Sex + Smoking status +WBCs + 30 control probe PCs (n=998)***

We further conducted principal component analysis on the residuals obtained from above step, and the resulting first five principal components could reflect unknown confounding effects.

***DNAme ∼ Age + Sex + Smoking status +WBCs + 30 control probe PCs (only for GDPH) + 5 PCs of residuals (n=998)***

***2)*** Each ESRD group comprised patients with end-stage renal disease resulting from a specific primary kidney disease, along with 196 shared controls. These ESRD groups included diabetic nephropathy (DKD-ESRD group, n = 538), polycystic kidney disease (PKD-ESRD group, n = 361), IgA nephropathy (IgAN-ESRD group, n = 340), hypertensive kidney disease (HKD-ESRD group, n = 277), and lupus nephropathy (LN-ESRD group, n = 266). The association between DNA methylation and ESRD group was evaluated using logistic regression models adjusted for potential confounders, including age, sex, white blood cells (WBCs) count, and smoking status. The outcome variable was ESRD attributed to specific primary kidney diseases, and the predictor of interest was the residuals of DNA methylation levels at individual CpG sites. Finally, we adjusted the test statistics for potential bias and inflation by employing a Bayesian-based method called “bacon” in each ESRD group^27^.

***ESRD in specific primary disease∼ DNAme residuals + Age + Sex + WBCs + Smoking status***

### Definition of primary kidney disease shared ESRD DMLs

Our goal was to identify ESRD-associated CpG sites shared across the five primary kidney disease groups. A CpG was defined as “shared” if it (1) achieved epigenome-wide significance (p < 1.29 × 10⁻⁸) in at least one ESRD group, based on Bonferroni correction for 774,631 CpG sites across five groups, and (2) showed Bonferroni-adjusted significance (p < 0.05/n) in the remaining four groups, where n is the number of significant CpGs in the original group (e.g., DKD-ESRD: 28 CpGs, p < 0.05/28; PKD-ESRD: 72, IgAN-ESRD: 58, HKD-ESRD: 13, LN-ESRD: 5). Applying these criteria, a total of 60 unique CpGs were identified.

To evaluate cross-ESRD group consistency, the heterogeneity of ESRD-associated CpG sites across different primary kidney diseases was assessed using Cochran’s Q test, as implemented in the “metafor” R package (v3.4.0). Statistical significance for heterogeneity was defined as a Bonferroni-adjusted p value < 0.05, based on the number of CpG sites tested.

Epigenome-wide ESRD associations were validated in KTPH (n=737) via CpG-level logistic regression with consistent adjustment for sex, age, smoking status, leukocyte fractions, and control probe PCs. Statistical significance in the validation cohort was determined at an FDR < 0.05 based on the number of CpGs tested. For each CpG site, we reported the beta coefficient from the logistic regression model, which represents the log odds ratio of ESRD per unit increase in residual methylation level, along with the corresponding standard error.

To evaluate potential confounding effects of key metabolic biomarkers, we implemented sequential sensitivity EWAS adjusting individually for: serum HDL-C, LDL-C, TC, TG, UA, and albumin levels. These analyses were restricted to participants with complete data for the respective biomarker in GDPH study, including 762 patients with available HDL-C measurements, 782 with LDL-C, 801 with TC, 791 with TG, 838 with UA, and 834 with albumin.

### Correlation analysis

To examine the variability of ESRD-associated DMLs across the five primary diseases, pairwise Pearson’s correlation coefficients were computed using the effect-size estimates (β) for the ESRD group in the discovery cohort. For each DML, effect sizes were calculated within each groups defined by the primary disease. Pairwise correlations were then computed between all possible pairs of primary disease-ESRD groups (e.g., DKD-ESRD vs. HKD-ESRD, DKD-ESRD vs. IgAN-DKD, etc.) to assess the consistency of methylation changes across etiologies.

To evaluate the consistency of effect sizes for ESRD-associated DMLs between the discovery and validation cohorts, Pearson correlation coefficients were calculated for the ESRD group. For each DML, effect-size estimates (β) from the discovery cohort were compared with those from the validation cohort.

### Trait enrichment analyses

We evaluated enrichment of ESRD-associated CpGs among trait-related methylation CpG sites in EWAS atlas^93^ and the MRC-IEU EWAS Catalog^94^, defining significant enrichment at Bonferroni-adjusted P value < 0.05 per trait. We restricted our comparative analysis specifically to EPIC array-based EWAS conducted in blood-derived tissues, to ensure comprehensive coverage of the 52 ESRD-associated loci.

We applied a permutation-based approach to characterize each ESRD-associated CpGS by generating a matched background set of 52 CpG sites with comparable methylation characteristics in blood samples. The permutation criteria included: (1) similarity of methylation means, (2) similarity of standard deviations (SD), and (3) a genomic distance greater than 5 kb from the ESRD-associated DMLs (to ensure independence). Specifically, for each ESRD-associated DMLs, we established thresholds using an adaptive sliding scale starting with a mean difference of 0.05 and an SD difference of 0.02, incrementally relaxing by 0.002 for mean and 0.005 for SD until at least 500 suitable permuted CpGs were identified. This adaptive threshold method was chosen because a fixed, strict cutoff did not yield sufficient permutations for some ESRD-associated DMLs, whereas a higher fixed cutoff was overly permissive for others. To quantify enrichment for each trait, the enrichment fold (EF) was calculated by comparing the number of ESRD-associated DMLs overlapping CpG sites linked to the trait (FDR < 0.05) with the average number of overlaps observed in 500 randomly selected CpG sets of the same size. We assessed enrichment significance by performing an exact binomial test, comparing observed overlap frequencies to those expected by chance. Bonferroni correction was applied to adjust the resulting P-values for multiple trait comparisons.

### Association analysis of complications in ESRD

We further evaluated the association of ESRD-associated DMLs with ESRD complications, such as anemia and CKD-MBD. CKD-MBD is primarily characterized by hypocalcemia, hyperphosphatemia, and high PTH levels. Uncontrolled anemia was defined as hemoglobin (Hb) levels below 115 g/L, based on the KDIGO Clinical Practice Guideline, which advise against the use of erythropoiesis-stimulating agents to maintain hemoglobin levels above 115 g/L in adults with CKD ^95^. Based on recommendations of the same guidelines^96^, it also recommends sustaining serum calcium and phosphate within physiological ranges, along with maintaining parathyroid hormone (PTH) at 2-9 times the assay-specific upper reference limit (approximately 130-600 pg/mL). Therefore, we examined the relationship of uncontrolled anemia(<115g/L), hypocalcemia (<2.1mmol/L), hyperphosphatemia (> 1.4mmol/L), elevated PTH levels (> 600pg/mL) and ESRD-associated DMLs using logistic regression models respectively. Serum calcium, phosphate, and PTH levels were measured using automated biochemical analyzers, and Hb levels were assessed via complete blood count analysis.

The analysis was restricted to ESRD cases with complete data on serum calcium, phosphate, parathyroid hormone (PTH), and hemoglobin (Hb) levels. Among ESRD patients in the GDPH cohort, 82.57% (n= 654) exhibited uncontrolled anemia (Hb < 115 g/L). Hypocalcemia was observed in 47.1% (n=645), hyperphosphatemia in 64.6% (n=658), and elevated PTH levels indicative of hyperparathyroidism in 22.4% (n=362). We used a Bonferroni-corrected P-value cutoff of 9.62 × 10⁻⁴, derived from dividing 0.05 by 52 tests, to address multiple comparisons.

### Genotyping procedures, QC, and imputation

Genomic DNA extraction from whole blood employed AxyPrep™ kits (Axygen AP-MN-BL-GDNA-250), ELISA-quantified (SpectraMax Plus 384). DNA samples meeting QC thresholds were genotyped on Illumina ASA BeadChips (ASAMD-24v1-0) per Infinium™ standards. Normalized array data underwent variant calling in GenomeStudio using Illumina’s embedded algorithms.

Pre-imputation QC of 924 genotyped individuals was conducted following established protocols. Samples with low call rates (<90%) or inconsistent sex information were excluded. Variants were filtered based on low call rate (< 98%), not conforming to Hardy–Weinberg equilibrium (*P* < 1×10⁻⁶), and insufficient minor allele frequency (MAF < 0.01). The final dataset included 338,114 autosomal variants. The genome coordinates of the build 37 (hg19) genotypes were converted to their equivalent build 38 (hg38) coordinates using the Genome Analysis Toolkit (GATK) LiftoverVcf tool, and and variants without unique mapping were filtered out.

We utilized the ChinaMap Imputation Server to impute autosomal single nucleotide polymorphisms (SNPs), aligning the data to the ChinaMAP.phase1.v1 reference panel^97^. Genotype imputation was conducted using Eagle2 for haplotype pre-phasing, followed by Minimac4 for imputation. Post-imputation quality control excluded variants with Hardy–Weinberg equilibrium P-values less than 1 × 10⁻⁶, INFO score at or below 0.6, or minor allele frequency below 0.01. After quality control, a total of 6,637,453 autosomal SNPs were retained for downstream analysis. To account for population stratification, principal component analysis (PCA) was performed using PLINK v1.9 ^98^.

### Identification of meQTLs

Using data from 924 individuals with genotype and methylation measurements, meQTL analysis was performed. Variants within a ±1 Mb region of each ESRD-associated differentially methylated loci were defined as cis-acting meQTL SNPs. Methylation M-values were adjusted using a linear regression model incorporating multiple covariates, including the top 30 principal components (PCs) from control probe intensities, age, sex, estimated blood cell fractions (NK cells, B cells, CD8 T cells, CD4 T cells, and monocytes) ^91^, the top 30 PCs from control probe intensities, the top 10 genotype PCs, and the top 5 PCs derived from methylation data. We then performed association testing between residual DNA methylation levels of ESRD-related DML, and SNP dosages using MatrixEQTL^99^, retaining SNP–CpG pairs that reached genome-wide significance (P < 5e-8).

To identify independent meQTL SNPs for each ESRD-related DML, we applied the conditional and joint analysis (COJO) and stepwise selection (COJO-SLCT) functions implemented in GCTA^100^. This method identifies conditionally independent variants through a stepwise forward selection procedure. A collinearity threshold set at 0.9 was consistently applied in analysis. Associations meeting the whole genome-wide significance (*P* < 5e-8) after conditioning were identified as independent signals. In addition, transcription factors (TFs) whose binding sites could potentially be altered by cis-meQTL SNPs associated with each ESRD-related CpG site were identified using snp2tfbs (http://ccg.vital-it.ch/snp2tfbs/)^101^.

### Association with eGFRcrea

To elucidate the potential of ESRD-associated DMLs for predicting the risk of progression to ESRD, we investigated the relationship between the DMLs and kidney function in CKD stages 1-3 by conducting a lookup analysis of 52 shared ESRD-associated DMLs using previously published human whole-blood EWAS data for eGFRcrea in populations with early CKD (stage 1-3) or healthy individuals. In most previous eGFR EWAS, eGFR based on serum creatinine concentration (eGFRcrea) is commonly used. Compared to ESRD patients, who typically have a mean eGFR value of <15 mL/min/1.73 m² (CKD stage 5), the included eGFRcrea EWAS datasets involved populations with mean eGFR values ranging from 49 to 91 mL/min/1.73 m², corresponding to CKD stages 1–3.

The studies included were: 1) Pascal et al. 2021^51^, including a discovery cohort with eGFR mean values of 87.4 ± 19.4 mL/min/1.73 m² and a replication cohort with eGFR mean values of 91.2 ± 20.3 mL/min/1.73 m²; 2) Li et al. 2023^22^, with eGFRcrea mean values of 80.6 ± 25.0 mL/min/1.73 m². All reported eGFR value ranges represent 95% confidence interval for the mean, reflecting the variability and distribution of renal function among study participants. Collectively, these data indicate that the majority of study participants had preserved to moderately impaired kidney function (eGFR mean value > 60 mL/min/1.73 m²), highlighting research focus on early stages of CKD.

### Colocalization analysis of loci associated with kidney function (eGFRcys-crea and BUN)

We applied colocalization analysis to investigate the same variants that influence both ESRD-related DMLs and kidney function. The meQTL data were derived from our study, while glomerular filtration rate based on cystatin C and serum creatinine (eGFRcys-crea) GWAS summary statistics data^102^ and BUN GWAS summary statistics data^103^ were downloaded from NHGRI-EBI GWAS catalog^104^. The eGFRcys-crea, which incorporates both eGFRcr and eGFRcys markers, provides greater accuracy than using either eGFRcr or eGFRcys individually. Moreover, BUN levels are linked to adverse renal outcomes, independent of eGFR, with a more significant impact on kidney disease progression, especially in the early stages of CKD.^105^ Therefore, these two markers can serve as complementary indicators for assessing renal function.

The eGFRcys-crea GWAS dataset comprises 406,504 samples, with a mean eGFRcys-crea value of 89.98 ± 14.158 mL/min/1.73 m², corresponding to early CKD. The BUN GWAS dataset comprises 635,969 samples. For each genome-wide significant SNP independently associated with an ESRD DML (meQTL SNP), the variants within a ±500 kb window were considered for colocalization analysis. Colocalization analyses were performed using the “coloc.abf” function from the “coloc” (version 5.2.3) package in R^106^, focusing on overlapping SNPs between the GWAS and meQTL datasets. Given that the array captures only 2–3% of CpG sites across the epigenome, some biologically relevant CpGs associated with ESRD may be missed. To address this limitation, colocalization was inferred when the posterior probability for either PP3 (shared locus with distinct causal variants) or PP4 (shared causal variant) exceeded 0.6, indicating evidence of colocalization.

### Mendelian randomization analysis

To assess causality, we applied Mendelian randomization to examine whether methylation at the identified CpG sites influences eGFRcys-crea and blood urea nitrogen (BUN) levels.

We applied a two-sample MR approach, “which is appropriate when exposure and outcome data are derived from different cohorts^107^. Methylation quantitative trait loci (meQTL) data (n = 923) from the current study were used. Independent genetic instruments for methylation levels were selected by LD clumping of cis-meQTL SNPs (*P*≤5 × 10⁻⁴) identified in our study. We assessed instrument validity by calculating the strength of genetic instruments, retaining only those with an F-statistic greater than 10 to ensure instrument validity. eGFRcys-crea GWAS summary statistics data^102^ and BUN GWAS summary statistics data^103^ were downloaded from NHGRI-EBI GWAS catalog^104^. Causal effect for individual CpG site was estimated using the Wald ratio method if only one instrumental SNP was available; when two or more instruments were present, the inverse-variance weighted (IVW) method was used. Both methods were conducted using the TwoSampleMR R package^108^.

### Colocalization analysis of gene-expression quantitative trait loci

We investigated whether ESRD-associated DMLs and gene expression share common causal variants by performing colocalization analysis between methylation quantitative trait loci (meQTLs) of 52 ESRD-associated DMLs and gene expression quantitative trait loci (eQTLs). Whole blood eQTL summary statistics were obtained from the eQTLGen Consortium, comprising 31,684 individuals ^109^. Blood-derived eQTLGen data in SMR format were converted to a readable summary statistic format for downstream analyses. Annotation was performed to convert these Ensembl IDs of eQTLGEN to human-readable gene symbols, using the R package “ChIPseeker” (version 1.42.0).^110^

For each genome-wide significant meQTL SNP independently associated with an ESRD-related DML, the nearest annotated genes within a ±500 kb window were considered for colocalization analysis. Colocalization analyses were performed using the “coloc.abf” function from the “coloc” (version 5.2.3)^106^ package in R, focusing on overlapping SNPs between the eQTL and meQTL datasets.

Bayesian posterior probabilities (PP) were estimated for five competing hypotheses: H0, no association in either the ESRD-related DML or the gene; H1, association in the DML only; H2, association in the gene only; H3, association in both the DML and the gene but driven by distinct causal variants; and H4, association in both the DML and the gene driven by a shared causal variant. Colocalization signals were identified by posterior probabilities greater than 0.8 in PP3 or PP4, reflecting shared genetic basis.

### Link CpGs to the target gene

Genes linked to methylated sites can be categorized into four main groups: those directly containing differentially methylated loci (DMLs), those regulated by DML-containing active enhancers in blood, those whose expression correlates with ESRD DML methylation levels in blood or kidney tissues and those located at the same genomic position as the corresponding cis-meQTL. To pinpoint the most probable target gene of ESRD-related DMLs, we employed a comprehensive multi-faceted approach (Supplementary Table 10).

The first group are the nearest genes, defined as those genes whose coding regions directly harbor the DMLs. Nearest genes to each DML were annotated based on the Infinium MethylationEPIC v1.0 B4 Manifest, which incorporates genomic annotations from both UCSC RefGene and GENCODE v12 databases. These annotations include CpG sites located within gene bodies or regions within 1500 bp upstream of transcription start sites (TSS), with a preference for overlaps in promoter regions over other genomic features (prioritized in the order: promoter, 5ʹ/3ʹ UTR, gene body, intergenic).

The second group comprised genes regulated by active enhancers in blood that harbor DMLs. Functional annotation was performed using a combination of genomic resources, including enhancer–gene connection datasets, three-dimensional (3D) chromatin interaction maps, and single-cell chromatin accessibility profiles. Enhancer-gene connections was mapped from two datasets in blood-related tissue: Epimap^111^ and Activity-By-Contact^112^, which inferred target gene by using a quantitative combination of enhancer activity and 3D contact frequencies (ABC model) or correlating the active enhancer marks with gene activity levels (Epimap). Promoter capture Hi-C (PCHi-C) maps provided information of genome-wide physical interaction between distal regulatory elements and promoters in 17 blood cell types^113^. Data from single-cell ATAC-seq (sc-ATAC seq)^114^ of peripheral blood mononuclear cells (PBMC) using the Cicero method was used to map DMLs to target genes based on significant co-accessible pairs of DNA elements.

The third group comprised genes whose transcriptional activity is influenced by DNA methylation levels. Functional annotation was performed using previously published expression quantitative trait methylation (eQTM) datasets derived from kidney or blood^11,32,115,116^.

The fourth group comprised genes whose cis-eQTL shared genomic loci with the cis-meQTL. We identified eGene-meCpG pairs through co-localization analysis with “coloc” (PP.H3 or PP.H4 ≥ 0.8). Results for cis SNP-expression (cis-eQTL) associations were obtained from eQTLgen^109^, while cis SNP-methylation (cis-meQTL) associations were reported in the current study.

For each annotation method, individual DMLs were linked to their most confidently associated target genes based on the high supporting evidence scores, including genes identified by the Activity-by-Contact maximum (ABC-max) prediction, Cicero gene activity scores (≥ 0.45), significant eQTM associations (FDR < 0.05), high-confidence chromatin interactions derived from Hi-C data (interaction frequency probability ≥ 0.78), EpiMap correlation scores (≥ 0.3), and posterior colocalization probability of eGene-meCpG pairs ≥ 0.8. The integration of these complementary databases enables a more comprehensive identification of genes influenced by ESRD-associated CpG sites.

### Gene Set Enrichment Analysis

Gene set enrichment analysis was performed using g:Profiler to identify biological pathways/process (https://biit.cs.ut.ee/gprofiler/gost)^117^. This tool incorporated multiple curated pathway databases, including Gene Ontology and KEGG, Reactome, and WikiPathways. The significance of pathway enrichment was assessed via the hypergeometric test, followed by Bonferroni correction to account for multiple comparisons. Pathways meeting the corrected significance threshold were considered significantly enriched.

## Data Availability

All summary statistics from the EWAS have been submitted to the OMIX platform hosted by the China National Center for Bioinformation (accession OMIX007372).

https://ngdc.cncb.ac.cn/omix

## Acknowledgements

We appreciate the involvement of all individuals who took part in this research.

## Ethics declarations

The research ethics of this project received formal approval from the Institutional Review Board of Guangdong Provincial People’s Hospital.

## Author contributions

Conceptualization was led by J.J.L, X.Q.Y, and L.S.C. Data analysis were performed by X.H.Z and J.J.X. Manuscript drafting was done by X.H.Z, D.C.S and J.J.X. Sample ascertainment and data generation were carried out by D.C.S, M.W, D.Y.F, W.C, Z.M.Y, Y.N.W, J.H.Z, H.G, P.F, X.J.T, J.H.C, Y.Z.K, and H.Q.Z. Clinical data collection and organization were conducted by D.C.S and J.J.X. Supervision was provided by J.J.L and X.Q.Y. Manuscript revision was performed by J.J.L, X.H.Z, D.C.S, J.J.X, X.Q.Y, L.S.C, Rajkumar Dorajoo, L.W, Resham Lal Gurung, and M. Yiamunaa.

## Code and Data availability

All summary statistics from the EWAS have been submitted to the OMIX platform hosted by the China National Center for Bioinformation (accession OMIX007372). Kidney disease-associated EWAS data were obtained from two public resources: the MRC IEU EWAS Catalog (http://www.ewascatalog.org/) and the EWAS Atlas (https://ngdc.cncb.ac.cn/ewas/atlas/index). eGFRcrea EWAS were obtaind from Pascal et al. 2021 (PMID 34887417) and Li et al. 2023 (PMID 37188670). The GWAS summary statistics for eGFRcys-crea and BUN, employed in colocalization and MR analyses, were retrieved from the GWAS Catalog (https://www.ebi.ac.uk/gwas/home), corresponding to the findings of Gabriel et al. (PMID: 39256582) and Verma A et al. (PMID: 39024449). In accordance with data privacy requirements, the individual-level data generated in this study are not publicly released. All processed datasets supporting this study key findings are available within the manuscript and Supplementary Materials. The code used for this study is accessible via <u>GitHub repository</u>.

## Supplementary Materials

Supplementary Figures 1–7 are included in this PDF.

Supplementary Tables 1–17 are provided as separate files to further support and elaborate on the findings.

