## Supplementary Figure 1 - 8 for "Genome-wide DNA methylation analysis revealed epigenetic mechanism underlying end stage renal disease"

### Supplementary Figures 1–7

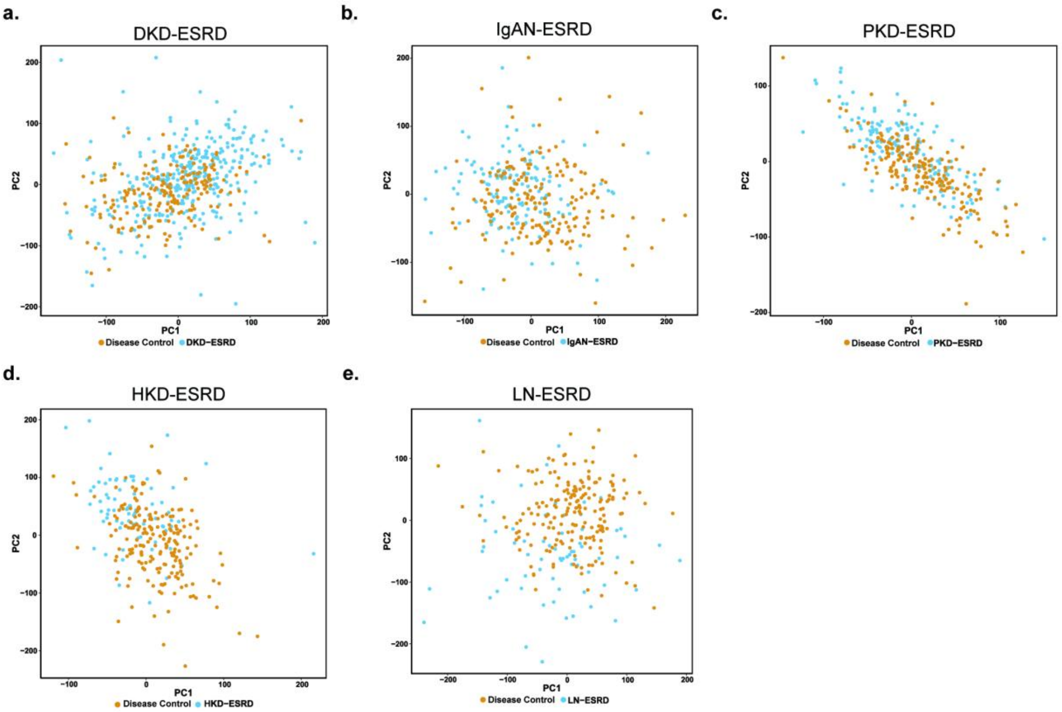


**Supplementary Figure 1. Principal component analysis of DNA methylation data adjusted technical batch effect and biological confounders in GDPH discovery cohort.**

Scatter plots of the first two principal components (PC1 and PC2) comparing ESRD groups caused by primary kidney diseases versus controls (n = 196): (a) DKD, (b) IgAN, (c) PKD, (d) HKD, and (e) LN. The same controls were used across all ESRD groups. DKD, diabetic nephropathy; PKD, polycystic kidney disease; IgAN, IgA nephropathy; HKD, hypertensive kidney disease; LN, lupus nephropathy.

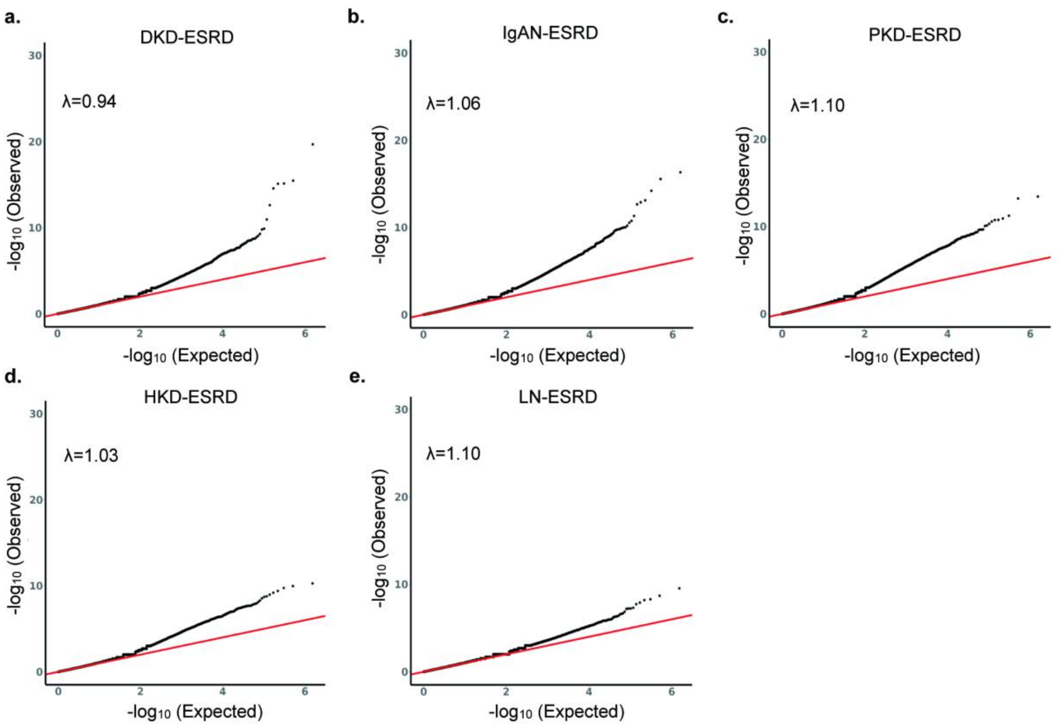


**Supplementary Figure 2. Quantile-Quantile plots displaying the distribution of observed $-\log_{10}(P)$ values relative to the expected null distribution.**

Epigenomic control inflation factors (λ), estimated using a Bayesian framework, indicate appropriate correction for technical bias across all analyses. EWAS analyses conducted for ESRD in five primary kidney diseases: **a)** DKD, **b)** IgAN, **c)** PKD, **d)** HKD and **e)** LN. The same controls (n = 196) were shared across ESRD in different primary kidney diseases. DKD, diabetic nephropathy; PKD, polycystic kidney disease; IgAN, IgA nephropathy; HKD, hypertensive kidney disease; LN, lupus nephropathy.

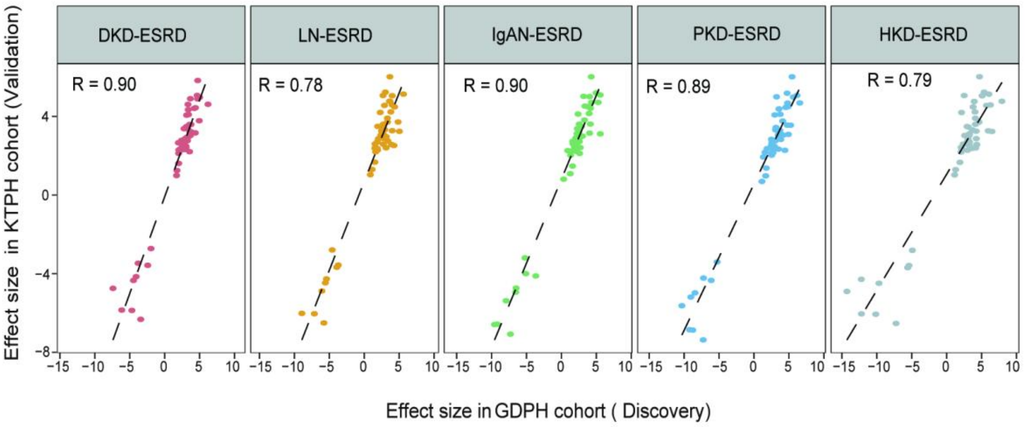


**Supplementary Figure 3. Relationship between Validation and Discovery Effect Sizes of 52 ESRD DMLs**

The Pearson correlation coefficient (r) is shown. DKD, diabetic nephropathy; PKD, polycystic kidney disease; IgAN, IgA nephropathy; HKD, hypertensive kidney disease; LN, lupus nephropathy.

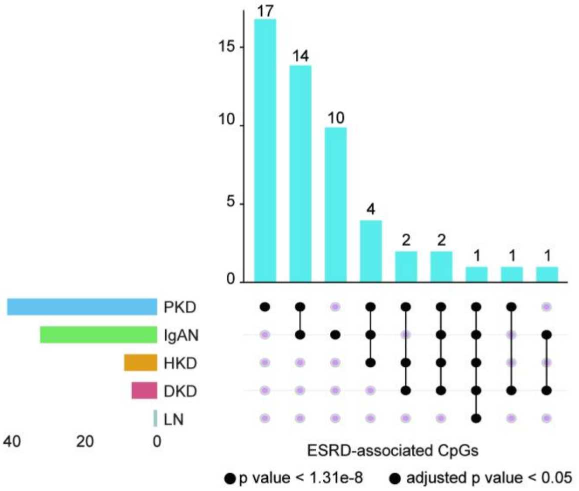


**Supplementary Figure 4. Intersection of common ESRD-associated CpGs across ESRD resulting from distinct primary kidney disease.**

ESRD in five primary kidney diseases: a) DKD, b) IgAN, c) PKD, d) HKD and e) LN.

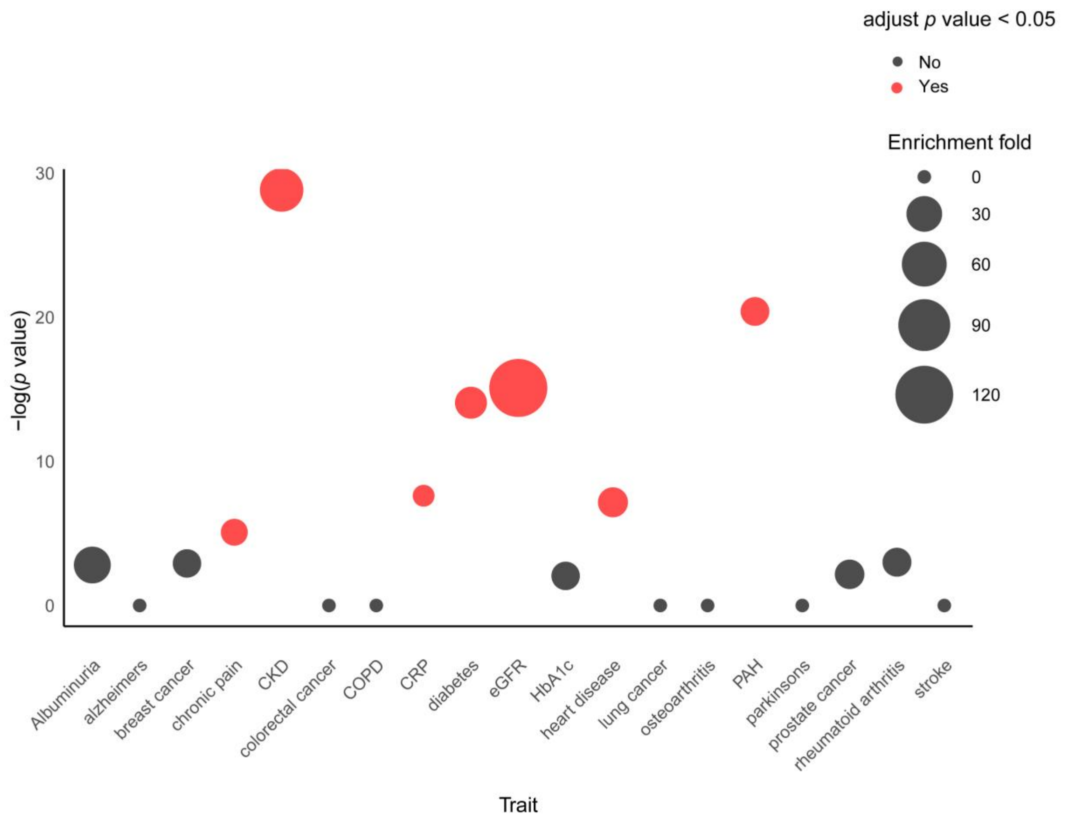


**Supplementary Figure 5. Disease enrichment analysis of 52 common ESRD-associated CpGs across ESRD groups.**

The x-axis represents different disease traits, and the y-axis shows enrichment significance as $-\log_{10}(P)$. Bubble size indicates the fold enrichment of DMPs for each trait. Bubble colors represent statistical significance: red for significant enrichment (Bonferroni-corrected p-value < 0.05) and gray for non-significant enrichment. P-values were calculated using a binomial test.CKD, chronic kidney disease; COPD, chronic obstructive pulmonary disease; hBA1C, Hemoglobin A1c; CRP, C-reactive protein; PAH, pulmonary arterial hypertension.

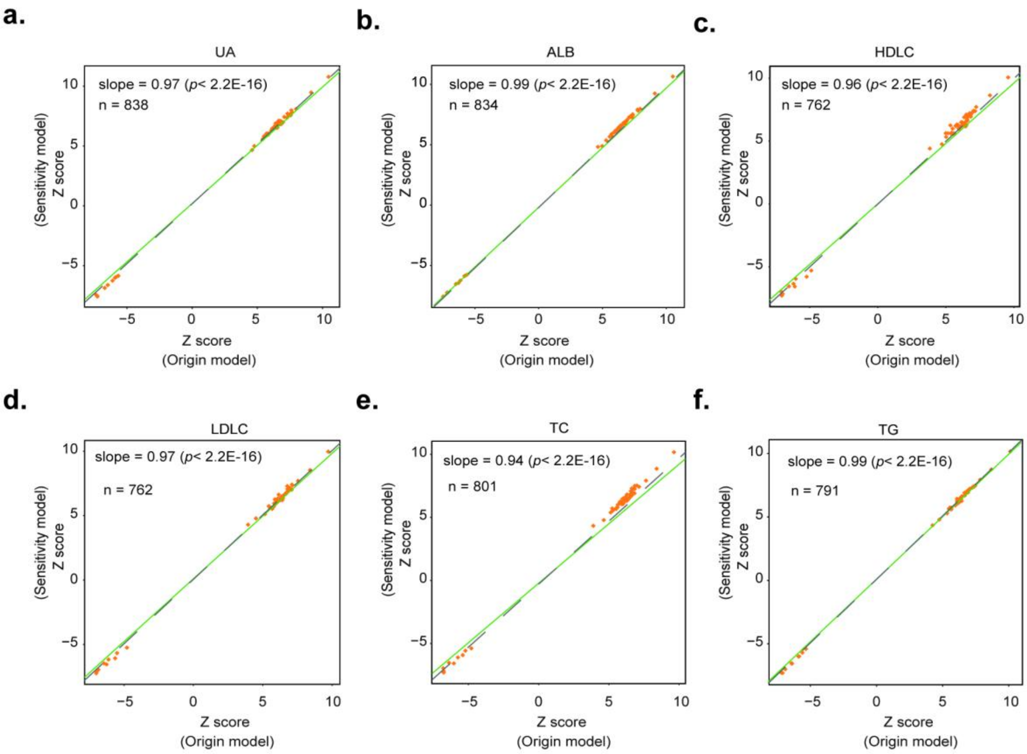


**Supplementary Figure 6. Scatter plots of EWAS sensitivity analysis in GDPH discovery.**

The original model was adjusted for age, sex, race, and confounders (control probe PCs and estimated leukocyte subtype proportions). The sensitivity model additionally adjusted for various clinical risk factors, including urine acid(UA), albumin(ALB), high-density lipoprotein cholesterol (HDLC), low-density lipoprotein cholesterol (LDLC), total cholesterol (TC), and triglycerides (TG).

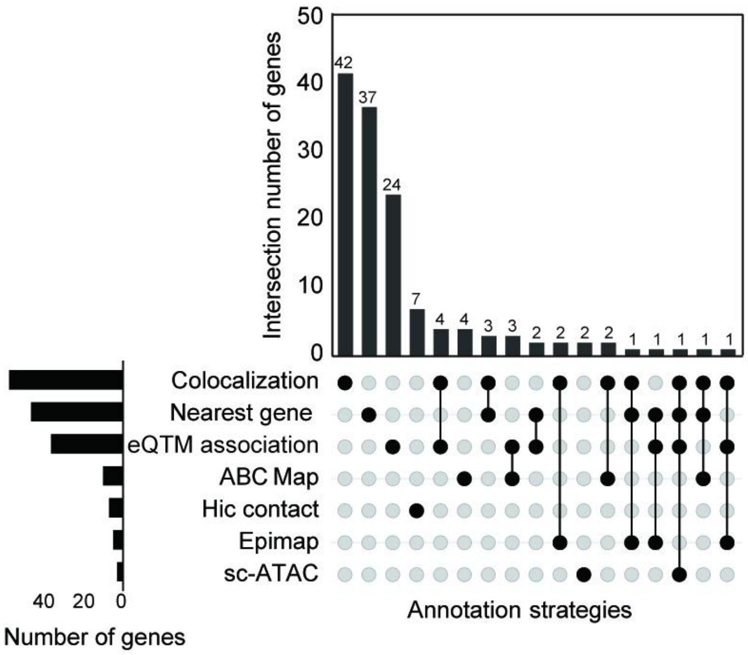


**Supplementary Figure 7. UpSet plot of candidate genes linked to ESRD-associated DMLs.**

Candidate genes were prioritized using multiple annotation strategies. The horizontal bars (bottom left) indicate the total number of genes identified by each strategy. The vertical bars (top) represent the number of genes uniquely or jointly annotated by different combinations of strategies, as indicated by filled and connected black circles below.
